# Cerebral oximetry monitoring versus usual care for extremely preterm infants: a detailed statistical analysis plan for the 2-year follow-up of the SafeBoosC-III randomised clinical trial

**DOI:** 10.1101/2024.12.09.24318704

**Authors:** Marie Isabel S Rasmussen, Mathias Lühr Hansen, Adelina Pellicer, Christian Gluud, Eugene Dempsey, Jonathan Mintzer, Simon Hyttel-Sørensen, Anne Marie Heuchan, Cornelia Hagmann, Ebru Ergenekon, Gabriel Dimitriou, Gerhard Pichler, Gunnar Naulaers, Jakub Tkaczyk, Hans Fuchs, Monica Fumagalli, Saudamini Nesargi, Siv Fredly, Tomasz Szczapa, Anne Mette Plomgaard, Bo Mølholm Hansen, Markus Harboe Olsen, Janus Christian Jakobsen, Gorm Greisen

## Abstract

**Background:** The SafeBoosC-III trial investigated treatment guided by cerebral oximetry monitoring for the first 72 hours after birth in extremely preterm infants and showed no effects on mortality or severe brain injury at 36 weeks’ postmenstrual age versus usual care. As severe brain injury in the neonatal period is not a strong predictor of long-term neurodevelopmental outcomes, the SafeBoosC-III follow-up study aims to assess the long-term benefits and harms of the experimental intervention versus usual care at two years of corrected age. This detailed statistical analysis plan outlines our approach for analysing outcomes in the SafeBoosC-III follow-up study.

**Methods:** The co-primary outcomes are 1) a composite of death or moderate-to-severe neurodevelopmental disability and 2) the mean Bayley-III/IV cognitive score. We will employ a 3-tier data model, incorporating routine clinical follow-up, parental questionnaires, and informal assessments to minimize missing data. All randomised participants with available data will be included in all analyses. Mixed-effect linear and logistic regression will be used to analyse the dichotomous and continuous co-primary outcomes, respectively. Sensitivity analyses will be conducted to address missing data and assess the robustness of our findings.

**Discussion:** The statistical analysis plan aims to ensure transparency and reduce the risk of outcome reporting bias. By including dichotomous and continuous co-primary outcomes, we aim to provide a comprehensive evaluation of the intervention’s effect on long-term benefits and harms.

## Background

Each year 50,000 children are born extremely prematurely in countries with routine neonatal intensive care. Although mortality rates are approximately 20%, survivors also face significant risks of neonatal brain injury and long-term neurodevelopmental disabilities (1). Long-term outcomes are essential considerations in neonatal trials as they provide insights into the efficacy and safety of interventions beyond the immediate neonatal period. However, achieving adequate follow-up is challenging due to factors such as relocation, socioeconomic constraints, and lack of awareness about the importance of follow-up assessments (2).

In neonatal cohort studies, loss to follow up typically varies from 25% to 50% but can be up to 70% when evaluating outcomes at two years (2). Studies indicate that parents of impaired children and those with a social disadvantage are less likely to participate in research (2-4). Missing data due to dropouts diminishes the statistical power of the trial and increases the risk of type II errors, hindering accurate assessment of intervention effects (5).

The SafeBoosC phase-II trial demonstrated that cerebral oximetry monitoring combined with a treatment guideline reduced the burden of cerebral hypoxia and hyperoxia by more than 50% within the first 72 hours after birth compared with blinded monitoring (6). While trends towards reduced mortality and severe brain injury were observed at 36 weeks’ postmenstrual age (PMA), subsequent follow-up at 24 months’ corrected age showed no significant differences in neurodevelopment between the two intervention groups (7). However, the phase II trial was not powered to detect a difference on clinical outcomes and therefore the larger SafeBoosC-III trial was planned as a multi-centre international trial investigating the clinical effects of cerebral oximetry-guided treatment in extremely preterm infants (8, 9). A total of 1,601 infants were randomised across 70 sites from Europe, the USA, China, and India. The hypothesis was that the intervention would reduce death or severe brain injury at 36 weeks’ postmenstrual age compared to usual care; but no significant differences were found (10).

Although severe brain injury diagnosed in the neonatal period is associated with long-term neurodevelopmental disability, this relationship remains strong only for the most severe brain injury (11). Assessment at 2 years’ corrected age is widely used both clinically and in research, which led to the design of the SafeBoosC-III follow-up study. The study protocol has been published (12). The present paper outlines the detailed statistical analysis plan for the long-term outcomes, including analysis principles, outcome definitions, methods for primary analysis and sensitivity analysis. This approach aims to reduce the risk of outcome reporting bias and data-driven analyses.

## Methods

The objective of the SafeBoosC-III follow-up study is to investigate the benefits and harms of treatment guided by cerebral oximetry monitoring in extremely preterm infants during the first 72 hours after birth, assessed at two years’ corrected age, as compared with usual care. It will be conducted according to the guidelines of the Helsinki Declaration (13). Details about the intervention and short-term outcomes (36 weeks’ PMA) have been reported elsewhere (8-10).

### Primary outcomes

The two co-primary outcomes are:

1. Death or moderate-or-severe neurodevelopmental disability (NDD) *(dichotomous outcome)*.

A child will be classified with moderate-or-severe NDD if any of the four following conditions are present:

- cerebral palsy with functional impairment corresponding to Gross Motor Function Classification Score (GMFCS) ≥ 2;
- a score below -2 standard deviations (SD) from the norm of a standardised developmental assessment (if using the Bayley-III/IV test, the cognitive score cut-off will be < 85), or an informal classification of moderate-or-severe NDD (for details, see below);
- vision impairment defined as moderately reduced vision, or only being able to perceive light or light reflecting objects; or blind in one eye with good vision in the contralateral eye;
- hearing impairment defined as hearing loss corrected with aids (usually moderate 40 to 70 decibels hearing level) or some hearing, but loss not corrected by aids (usually severe 70 to 90 decibels hearing level).
- Bayley-III/IV mean cognitive score *(continuous outcome)*.

### Exploratory outcomes

The exploratory outcomes are:

- Any daily medication for the last two months *(dichotomous outcome)*.
- Any other chronic illness (defined as any problem which has been diagnosed by a doctor and which is expected to last more than a few months; causes problems in everyday life; or is a risk of early death or disability) *(dichotomous outcome)*.
- Mean head circumference in cm *(continuous outcome)*.
- Mean height in cm *(continuous outcome)*.
- Mean body weight in kilograms, with one decimal *(continuous outcome)*.
- Hospitalisation since being discharged home from the index hospital admission, based on parental report *(dichotomous outcome)*.
- Parent Report of Children’s Abilities-Revised (PARCA-R) non verbal cognitive mean score (*continuous outcome)*.
- Parental report of the child’s thriving *(dichotomous outcome)*.
- Parental report of worries regarding the child *(dichotomous outcome)*.

The component of the dichotomous co-primary outcomes will be reported individually for the two groups separately, including effect estimates with confidence intervals and will be taken into consideration when interpreting the results.

### Power estimations for the co-primary outcomes

The sample size calculation of 1,600 participants for the SafeBoosC-III trial was based on mortality and the prevalence of survivors with severe brain injury in the SafeBoosC-II trial, i.e. a 22% relative risk reduction in the composite primary outcome from 34% to 26.5% (absolute risk reduction of 7,5%), at an alpha level of 5%, and a power of 90% (8). Of the 1601 participants randomised, 1276 were alive at 36 weeks’ post menstrual age and would potentially be available for assessment in SafeBoosC-III follow up study. The following calculations were carried out using the software OpenEpi (Dean AG, Sullivan KM, Soe MM. OpenEpi: Open Source Epidemiologic Statistics for Public Health, Version 2.3.1.)

Based on results from two previous randomised clinical trials investigating neuroprotection in preterm infants (14, 15), it was estimated that the proportion of children with the outcome death or moderate-or-severe NDD would be 50% in the usual care group. An indicative power calculation showed that if we wanted to test an absolute risk difference of 8%, between the cerebral oximetry and usual care group, at an alpha of 2.5% and a sample size of 800 participants in each group, i.e. a total of 1600, we would reach a power of 80% for this outcome. As the study progressed, some sites were excluded or withdrew, and missing data was anticipated. An updated power calculation shows that a realistic estimate would be to have 650 participants in each group, i.e. a total of 1300, and with the same absolute risk difference and alpha, we can thereby reach a power of 75%.

Based on answers to a questionnaire on systematic routine follow up, it was expected that 65% of the sites would be able to provide data from a Bayley-III/IV assessment around two years’ corrected age for each child. Assuming these sites would enroll their proportion of the eligible participants in the SafeBoosC-III trial, approximately 850 participants would be available for Bayley-III/IV assessments at two years of age.

An indicative power calculation showed, that if we wanted to test a mean difference of five points on the Bayley cognitive score, with a standard deviation of 20 (Cohens d 0.25) between the cerebral oximetry and the usual care group, at a 2.5% alpha, with a sample size of 425 participants from each group in the follow up study i.e. a total of 850 participants, we would reach a power of 90%. In regard to the same factors as mentioned in the above, the updated power calculation shows that, a realistic estimate would be to have 250 participants in each group, i.e. a total of 500, and with the same absolute mean difference and alpha, we can thereby reach a power of 71%.

### Assessment of the dichotomous co-primary outcome

We will use three tiers of data to assess death or moderate-or-severe neurodevelopmental disability: 1) routine clinical data from healthcare records; 2) parental questionnaire; and 3) informal assessments of neurodevelopment. Data from all three tiers will be used for the primary analysis to minimize missingness.

#### Tier 1 (top priority)

use of clinical records with formal developmental assessments when the child is age 18-30 months of corrected age. A child will count as an event if it has been diagnosed with either cerebral palsy, visual impairment, or hearing impairment, according to the definitions described previously. They will also count as an event if the child has a Bayley-III cognitive score below 85, as done in previous trials and recommended by experts (16). If the child does not have a Bayley score, the results of other neurodevelopmental tests will be used. Many tests are used as part of routine follow up in the SafeBoosC-III follow up study, but not all are considered a cognitive/neurodevelopmental assessment. Therefore, prior to data analysis the used assessments were reviewed, and we chose to consider the following assessments valid as a cognitive measure: Bayley II, Griffiths III, Gessell, Ages and Stages Questionnaire, Denver Developmental Screening, Peabody, Developmental Profile 3, Battelle Developmental Inventory, Brunet Lezine, Haizea-Llevant or a Schedule of Growing Skills (in prioritised our order). Other tests will not be used for the assessment of neurodevelopmental disability. The classification of each participant will be provided by the blinded assessor, who will assess all available data, possibly including locally used standards, and decide whether the score is indicative of moderate-or-severe neurodevelopmental disability or not.

#### Tier 2 (second priority)

if the blinded assessor is unable to classify the child on any of the above tier 1 criteria, or data are outside the 18–30-month window, data from the parental questionnaire will be used instead. Although the 30-month upper limit was set to define a timeframe, the validity of clinical data generally improves over time. Therefore, children just beyond this upper limit would still be included. The parents are asked if their child has been diagnosed with cerebral palsy, or if the child was not able to walk independently at 2 years of corrected age; if their child is blind in one or two eyes or has poor vision even with glasses; or if their child has hearing aids or cochlear implants. “Yes” to any of these questions will count as an event in either cerebral palsy, visual-or hearing impairment. Furthermore, if no neurodevelopmental tests have been performed, this score will be substituted by the score of the PARCA-R non-verbal cognitive score according to standardisation. All questionnaire data may be substituted if data from clinical follow up is not available assessment of the outcome moderate-or-severe NDD.

#### Tier 3 (third priority)

if data from tier 1 and 2 are incomplete, the blinded assessor will make an informal assessment based on all clinical records from when the child was 12 months and forward to complete the criteria for classification of the child as moderate-or-severe NDD or not. An informal assessment can be made for the entire classification of the child or only for the cognitive measure.

### Explanatory variables

Additional clinical data on trial participants will be collected from clinical records, to compare characteristics between the two groups. These data will be presented in a table in the main publication. Tests of statistical significance will not be undertaken for explanatory variables. Categorical data will be summarised by numbers and percentages. Continuous data will be summarised by mean and 95% confidence interval (CI) if normally distributed or by median and interquartile range (IQR) if non-normally distributed.

### Central monitoring

A central monitoring plan was designed based on the monitoring plan from the SafeBoosC-III trial (17). The monitoring plan was divided into two parts: 1) missing data monitoring and 2) monitoring of quality deficiencies and deviations.

### Part 1 – *missing data monitoring*

For each site, missing data for both clinical data and parental questionnaires were reported. Data were considered missing if the “blinded 2-year follow-up” data entry was not completed by 24 months of corrected age plus one month. Although data were not technically missing until after 30 months of corrected age, this threshold served as a reminder to investigators to complete the data entry in a timely manner. Parental questionnaires, hosted on REDCap (18), were standardised to be filled out between 23.3-27.5 months of corrected age. The completion rates were reported for each site in the monthly newsletter circulated to all investigators and uploaded to ‘safeboosc.eu’. Identified study IDs with missing data were sent directly to the relevant primary investigators by the study manager.

### Part 2 *– quality deficiencies and deviations*

To identify sites with noteworthy data deviations, monitoring of data quality deficiencies was conducted when the last participant turned 24 months of corrected age and again at 30 months of corrected age. The focus was on identifying noteworthy data deviations such as outliers, systematic deviations due to misunderstandings, and suspected fabricated data. Quantitative measures were analysed for outliers. Systematic deviations were reviewed, such as above-average numbers of participants lost to follow-up or inconsistencies in cognitive assessment scores. Inputted dates for time of assessments were performed as well. Any identified issues prompted direct communication with investigators for verification or correction.

Additionally, data were examined for suspected fabrication by analysing unexpected distributions or variances in binary data.

The results from this monitoring were logged in a ‘central monitoring log’ by the monitoring group, ensuring that any issues were documented and addressed appropriately.

### Level of significance

Significance thresholds will be evaluated in adherence to a five-point procedure as described by Jakobsen and colleagues (19). First, the confidence intervals and p-values for all outcomes will be calculated. Secondly, the Bayes factor will be calculated for the two co-primary outcomes. A Bayes factor less than 0.1 has been chosen as significance threshold (20). By calculating the Bayes factor, it will be possible to interpret the results of the primary outcome in relation to former trial results. The confidence intervals will be presented as two-tailed. Superiority of the intervention will only be claimed, if at least one of the two co-primary outcomes is statistically significant. To correct for multiple testing, the threshold for statistical significance will undergo Bonferroni adjustment, and thus a p-value of 0.025 for each primary outcome is chosen. If statistical significance is shown, clinical significance will be assessed as well, by taking all possible benefits and harms into the consideration. As the exploratory outcomes are only hypothesis generating, and since the primary conclusion will be based on the primary outcome results, no adjustments will be made to the statistical significance threshold for the exploratory outcomes.

### Primary analyses

The primary analyses of all outcomes will be based on the intention-to-treat population. Complete case mixed-effect linear regression and mixed-effect logistic regression will be used to analyse the dichotomous and continuous co-primary outcomes, respectively. In the regression models, ‘site’ will be included as a random effect, allowing for variability across sites. Gestational age below or above 26 weeks and ‘group allocation’ will be included as fixed effects, since both were stratification variables for group allocation in the trial.

### Handling of missing data

We anticipate missing data due to varying levels of engagement in follow-up assessments among sites. Sites may withdraw from the follow-up study. Further, sites that do not actively pursue participation in the study may be excluded and will not count as missing data.

Missing data will be handled according to the recommendation by Jakobsen and colleagues (21). Assuming that data is missing at random, we will apply chained equation multiple imputation, to predict missing values based on observed data. Variables to be used for imputation of outcomes are gestational age, sex, respiratory treatment, intraventricular haemorrhage, retinopathy of prematurity, bronchopulmonary dysplasia and necrotizing enterocolitis. These variables are selected for their known association with childhood outcomes in cohorts of extremely preterm children and their availability in the dataset.

### Sensitivity analysis

We will perform several sensitivity analyses to assess the robustness of our findings. First, we will compare sites with high follow-up rates (>90%) to those with lower follow-up rates (<90%) to explore potential differences in effect estimates. Additionally, we will conduct a sensitivity analysis excluding the informal assessment of moderate-or-severe NDD to evaluate the impact of this exclusion on the overall results.

We will carry out a per-protocol sensitivity analysis, including only those participants who adhered strictly to the intervention protocol—defined as continuous cerebral oxygenation monitoring during the first 72 hours of life or until death, according to the SafeBoosC-III protocol (9). As in the SafeBoosC-III trial, we will also analyse the results using a random-effects meta-analysis (8). Additionally, we will perform an analysis including all PARCA-R scores, extrapolating data from questionnaires completed outside the standardized timeframe. Finally we will use standardised mean differences (SMD) to compare various developmental assessments at two years of age. We will be pooling scores and convert them to SMD, allowing for comprehensive evaluation of intervention effects across different assessment tools.

### Assessing potential bias due to missing data

We will quantify potential biases arising from missing data in the dichotomous co-primary outcome. We will calculate a ‘true’ relative risk, to account for potential bias when the rate of missing data differ between the randomisation groups and, at the same time differ between the outcome groups. We will use the observed outcome rates, the observed proportions of missing data in the intervention groups and introduce assumptions on the rates of missingness in the outcome groups, from 1:2 to 2:1 and random skewness in both directions corresponding to a p-value of 0.05 as calculated by the Chi-square test.

### Assessment of underlying statistical assumptions

We will systematically assess the statistical assumptions for each method, following recommendations by Nørskov and colleagues (22). Specifically, we will evaluate potential interactions between covariates and the intervention in all regression analyses. First-order interactions will be tested individually, and interactions will only be considered significant if they meet the Bonferroni-adjusted threshold (0.05 divided by the number of possible interactions) and show a clinically relevant effect. If a significant interaction is identified, we will present both a stratified analysis and an overall analysis including the interaction term in the model.

### Twins and their intra-cluster correlation

Approximately 30% of the extremely preterm population may be twins or triplets (23). In the SafeBoosC-III trial this proportion was 27% (10). This may pose as a problem in the statistical analysis, as the outcomes may correlate among twins and thereby diminish the effective sample size and thus, the statistical power (24). The SafeBoosC-III trial randomised multiple births to the same group allocation. In the SafeBoosC-II trial, the intra-class correlation coefficient (ICC) of the burden of hypoxia within twin pairs was negligible. Furthermore, a simulation study evaluating the potential effect of twin intra-cluster correlation in the SafeBoosC-III trial, revealed that even with twin proportion as high as 30% and relatively high ICC values (0.2), the statistical power and coverage of the confidence intervals would only be marginally affected (8). Therefore, the sample size of 1601 infants of the SafeBoosC-III trial was not adjusted for possible twin ICC. For the two primary outcomes of the follow-up study, the twin data will also be analysed as independent observations. However, we will conduct a sensitivity analysis taking into consideration, the twin ICC’s potential effect on the primary outcomes. This will be conducted using the generalised estimating equation approach utilising an exchangeable covariance matrix with site and stratification variables as fixed effects. Additionally, the ICC of the co-primary outcomes of this follow-up study will be calculated, reported and compared to the ICC of the SafeBoosC-III trial.

### Blinding of statisticians

All data managers, statisticians, and those responsible for drawing conclusions will be blinded to the group allocation to ensure unbiased analysis. Two blinded statisticians, connected to The Copenhagen Trial Unit, will independently perform all statistical analyses. Each statistician will conduct the analysis separately according to the present detailed statistical analysis plan, with their statistical reports published as supplemental material. As the statistician who conducted the 36-week analysis will be involved in the 2-year outcome analysis, additional blinding measures will be applied to prevent any potential bias. Specifically, the statistician will not have access to information linking previous data with current group allocations, and any prior analyses will be presented in a way that prevents revealing group identities.

### Publication plan

Once all data from follow-up assessments has been analysed, the results will be published in a peer-reviewed international journal. One author per participating site can obtain co-authorship and the blinded assessor completing the data entries will obtain non-byline co-authorship. Ancillary studies with results potentially affecting equipoise or blinding, shall not be published before the main results of the follow-up study have been published.

## Discussion

The SafeBoosC-III follow up study aims to investigate the benefits and harms of treatment guided by cerebral oximetry monitoring in extremely preterm infants during the first 72 hours after birth versus usual care assessed at outcomes at two years’ corrected age. This discussion focuses on methodological challenges with a focus on handling missing data and bias.

### Potential bias across outcome and randomisation group

Neonatal trials often struggle with maintaining complete follow-up due to factors such as relocation, socioeconomic constraints, and lack of awareness about the importance of follow-up. In perinatal and neonatal trials, nearly 90% report a significant amount of incomplete outcome data (25). In the SafeBoosC-III trial, missing data poses a significant threat. The impact of missing data is twofold: it decreases the representativeness of the sample, and without adequate handling, it may introduce bias. Bias is introduced if those lost to follow up have less favorable outcomes. If children with moderate or severe NDD are less likely to be included in the follow-up, the estimate of the relative risk will remain unaffected, if the proportion of missing data is similar in both groups. However, the absolute measures of effect will be biased downwards. This scenario is plausible if these children’s health care records are inaccessible to the blinded assessor or if their parents, who may already have extensive contact with the healthcare system, are less inclined to respond to questionnaires. Socioeconomic factors may exacerbate this bias (26). Conversely, children with favorable outcomes might also be underrepresented if they are discharged from routine follow-up, leading to decreased parental interest in participating. Nevertheless, this would still not impact the relative risk estimate if the pattern is similar in the two groups. We will conduct sensitivity analysis checking if data is missing at random or missing not at random and use relevant analysis, accordingly. Furthermore, a calculation on a ‘true’ relative risk which adjusts for missing data will be calculated, giving an estimate on how vulnerable the effect estimate is to bias.

It is unlikely that missing data is influenced by randomisation group as the intervention is only three days in the beginning of a hospital stay, which for survivors last several months. Further, the intervention is only part of a complex package of care. A significant proportion of parents who give written consent for a trial in the neonatal period do not later remember having done so (27).

### Mitigation measures

As the follow up study was not initially planned, some sites withdrew. Additionally, sites that did not actively pursue participation in the study were given six months to improve. If no progress was made, they were excluded after consultation with the national coordinator. As no data were collected from excluded sites and as randomisation was stratified by site, this should prevent bias to the relative risk estimate. While this improves data quality and interval validity, it reduces external validity, limiting generalisability. In particular, if sites contributing data to the follow-up study provide better care, or applied the intervention more carefully this could affect the estimates of absolute measures of effect. However, since the random-effects meta-analysis of site-specific effects showed minimal heterogeneity in the SafeBoosC-III trial, it is unlikely to impact the follow-up study results (10).

As regards the continuous co-primary outcome, many sites rely on formal neurodevelopmental assessments such as the Bayley-III. As not all sites use the Bayley-III, but instead use Bayley-II, Griffiths or Denver etc. loss to follow-up for this outcome will be much higher. A mixed-effect regression model will adjust for site-level differences and furthermore we will calculate the SMD by standardising the intervention effect across assessments.

A central monitoring plan was implemented to minimise missing data and improve data quality by addressing missing data and quality deficiencies in real time. Outliers and suspected data fabrication were addressed but no deviations were found. However, the systematic deviations proved impactful and served as a form of data cleaning which lead to enhancement of data quality. Earlier and continuous monitoring throughout the study may have further enhanced data quality. Additional strategies to limit loss to follow up have previously been published (12).

### Patient-relevance of the two-year outcome

In the SafeBoosC-III follow-up trial, we have chosen a two-year outcome balancing ability to become conducted, timeliness, and reasonable patient relevance. The two-year outcome is commonly used in neonatal trials for detecting severe impairments like cerebral palsy and cognitive delays. However, while its predictive value is good to the severe cases, it may overlook neuro-psychological disabilities that emerge later (28). The binary classification of moderate or severe neurodevelopmental disability, while practical, simplifies complex developmental patterns and may lack statistical power. To address this, a continuous measure in the form of the Bayley-III cognitive score is included. Such offer greater statistical precision but are more difficult to interpret clinically, as early developmental scores in the normal range only moderately correlate with long-term measures of cognition (29). The two co-primary outcomes were combined to strengthen both the statistical power and clinical relevance of the SafeBoosC-III follow up outcomes, as planning for a longer follow up appeared unrealistic due to attrition rates and funding constraints.

### Study status

The protocol is registered at clinicaltrials.gov NCT05134116. The first participant was followed up in September 2021 and anticipated date of study completion is October 2024. Recruitment status can be accessed at www.safeboosc.eu.

## Data Availability

Not applicable

https://safeboosc.eu/

## Consent for publication

Not applicable

## Availability of data and materials

Not applicable

## Author’s contributions

MSR, MLH, CG, JCJ, MHO and GG contributed to the conception and design of the manuscript, drafted the manuscript, and will give final approval of the version to be published. AP, ED, JM, SHS, AMH, CH, EE, GD, GP, GN, JT, HF, MF, SM, SF, TS, AMP, BMH revised the main manuscript critically for important intellectual content and will give final approval of the version to be published.

